# Smartphone app-based Integrated Cognitive Control Training for Anxiety Disorders: Study protocol for a Multi-site Randomized Control Trial

**DOI:** 10.64898/2026.02.05.26345626

**Authors:** Himani Kashyap, Snehil Gupta, Haroon Lone, Renuka Mulay, Ammu. G. Thampi, Srinivas Balachander, TS Jaisoorya, Paulomi M. Sudhir, K. Thennarasu, Vikas Menon, Triptish Bhatia, Smita Deshpande, Konasale Prasad, YC Janardhan Reddy

## Abstract

**Background:** Cognitive deficits in anxiety disorders (ADs) contribute to clinical and socio-occupational dysfunction, necessitating targeted interventions.

**Novelty:** Integrated Cognitive Control Training (ICCT), a novel intervention, has demonstrated benefits in other disorders, however, remains unexplored in ADs. With its process-specific training and multi-pronged exercises for stimulation, metacognitive training and generalization, it has potential for enhancing cognitive functions in ADs.

**Objectives:** This paper describes the study protocol for a multi-site randomized controlled trial (RCT) to test efficacy of ICCT in individuals with ADs.

**Methods:** Adults diagnosed with ADs (n=100) will be recruited across two sites. Following baseline assessments, they will be randomized to either ICCT (8 weekly sessions) or Treatment As Usual (TAU). ICCT will be delivered through once-weekly therapist-guided, and smartphone app-based (‘*Cogtrain’*) homework (20-30 mins, 4-6 times per week). Multimodal assessments will be carried out at baseline, mid-intervention (4 weeks), post-intervention (8 weeks) and follow-up (20-24 weeks). The primary measure comprises Hamilton Anxiety Rating Scale, with secondary measures of Work and Social Adjustment Scale (socio-occupational functioning), neuropsychological tests (attention, memory and executive functions) and functional Magnetic Resonance Imaging of the cognitive control circuits. Intervention feasibility and acceptance metrics (response rate, intervention relevance) will also be recorded. Quality assurance and ethical procedures will be documented.

**Expected outcome:** The ICCT is expected to enhance cognitive functioning in adults with ADs, in addition to symptom reduction, changes in underlying neural circuits of cognitive control and improve overall functioning. Digital delivery through a smartphone app may provide a cost-effective and scalable intervention, useful in resource-constrained settings.

**Key Messages:** This multi-site randomized controlled trial evaluates a novel, smartphone-delivered Integrated Cognitive Control Training (ICCT) program for adults with anxiety disorders, targeting core cognitive deficits that contribute to functional impairment. By combining therapist-guided sessions with app-based training and multimodal assessments, the study examines both clinical and neural outcomes. Findings are expected to inform the scalability and feasibility of process-based digital cognitive interventions for anxiety disorders, particularly in resource-limited settings.

**Protocol Registration:** ***Trial registry name***: Clinical Trial Registry of India

***URL***: https://ctri.nic.in/Clinicaltrials/**************

***Registration number***: CTRI/202*/**/******

## Introduction

### Background

Anxiety disorders (ADs), characterized by debilitating anxiety, include generalized anxiety disorder (GAD), social anxiety disorder (SAD), panic disorder (PD) and specific phobia.^1^ Affecting 2-3% of India’s population,^2^ ADs are among the top causes of disability,^3^ with treatment gaps upto 85%.^2^

Cognitive deficits in ADs, primarily cognitive control (CC) - comprising working memory(WM), set shifting, and inhibitory control,^4,5^ can predict development of anxiety symptoms up to nine years later^6^ and contribute to socio-occupational dysfunction and economic burden.^5^

### Literature review

Cognitive deficits and biases in anxiety are interlinked, underpinned by neural networks-Attention control theory proposes that anxiety impairs goal-directed attention, and increases stimulus- and threat-driven attention.^7,8^ Neuroimaging studies demonstrate hyperactivation of the amygdala and insula with hippocampal dysfunction,^9^ probably explaining memory dysfunction and information processing biases in ADs.

Enhanced CC can reduce amygdala activation and anxious responses to emotional stimuli.^10^ Studies suggest anxious individuals can suppress attentional biases by task-related effort,^8^ underscoring CC as a critical therapeutic target. Cognitive training (CT) is a promising method for enhancing neurocognitive functions, underlying neural mechanisms, and putatively, clinical recovery in neuropsychiatric conditions.^11^

Evidence for CT in ADs is mixed. In high trait–anxious adults, 3-week WM training improved attentional and inhibitory control, resting-state electroencephalogram, and reduced trait anxiety.^12^ Emotional WM training in SAD demonstrated reductions in peak anxiety and post-event processing.^13,14^ In contrast, adaptive WM training in anxious and depressed individuals did not improve WM capacity, rumination, or anxiety/depression symptoms.^15^ Such ‘narrow transfer of training’, with limited generalization to untrained domains, is largely attributable to methodological limitations, including restricted focus on single domain (mainly WM), and inadequate training dose or duration (1-10 sessions, 20-45 minutes).^16–19^

### Novelty

Integrated Cognitive Control Training (ICCT), a novel intervention, has demonstrated improvements in obsessive-compulsive disorder and depression, on CC and other untrained domains.^16,20–22^ ICCT is guided by evidence-based principles - focus on transdiagnostic construct of CC, using process-specific training,^23-25^ incorporating ‘practice standards’ - metacognitive strategy training, enhancing self-awareness, and generalization,^26,17,19^ adhering to recommended dose, duration and frequency,^18^ and delivery via smartphone app for scalability and generalization. Neural mechanisms will be examined with neuroimaging.

### Objectives

The primary objective is to compare ICCT and Treatment As Usual (TAU) on pre-mid-post intervention anxiety-score changes across 8 weeks. Secondary objectives are to compare groups on socio-occupational functioning, cognitive functions, haemodynamic correlates, and intervention and acceptance metrics (response rate, intervention relevance).

## Methodology

### Study Design/ Study Participants/ Setting

A randomized controlled multisite parallel trial design, with superiority framework will compare two intervention arms, ICCT+TAU and TAU, across two sites. Participants will be assessed at baseline (Week 1), midline (Week 5), post-intervention (Week 10-12) and follow-up (Weeks 20-24). All participants will continue to receive medication, psychoeducation and routine follow-up care.

All consecutive participants with a Diagnostic and Statistical Manual of Mental Disorders, Fifth Edition (DSM 5) diagnosis of ADs (GAD, SAD, PD or specific phobias), diagnosed using Diagnostic Interview For Anxiety, Mood, and Obsessive-Compulsive And Related Neuropsychiatric Disorders (DIAMOND)^27^, right-handed, aged between 18-50 years, with minimum 7th standard education, working knowledge of English, and access to a smartphone, will be recruited from inpatient and outpatient services at site A and outpatient services at site B (masked for review), India. In the three months preceding participation, participants are required to be stable on/off medication, with no exposure to neuromodulation, or < 3 sessions of structured psychotherapy, and no changes to treatment during the course of the study. Participants with a history of severe mental disorders (psychotic disorders and bipolar affective disorders), acquired brain injuries (e.g., epilepsy, tumours, cerebrovascular accidents, degenerative conditions, traumatic brain injury), comorbid substance/behaviour addiction except tobacco, and developmental disorders including intellectual disability, will be excluded. Individuals who attend 80% of training sessions will qualify as completers, while participants who complete baseline assessments and initiate treatment but later communicate unwillingness to attend, or miss three consecutive sessions will be considered as dropouts.

### Interventions

#### Integrated Cognitive Control Training

ICCT^16,20–22^ is based on the premise that improved cognitive control will enhance cognitive and socio-occupational functioning, symptom expression and neural circuits. It uses an integrated approach with techniques for cognitive stimulation using a smartphone app *(Cogtrain)*, metacognitive awareness and monitoring, and transfer/generalization to real-world activities (Fig 1).

Participants will receive 24 hours of total training - once-weekly sessions with a therapist (in-person or online, based on preference), along with homework practice (20-30 minutes 4-6 times per week). Using a range of app-based games (*Cogtrain*), participants will be trained in cognitive stimulation - exercises of attention, cognitive flexibility, planning, decision-making and memory. Metacognitive training will focus on understanding and regulating cognitive functions, use of appropriate strategies, and pruning of ineffective strategies. Generalization of training to participant’s daily activities will be done through discussions and role plays, and using cognitive control strategies to manage distractions and anxiety. Homework and app engagement metrics will be logged and reviewed through the app.

ICCT has been previously developed for OCD and depression. It will be adapted and piloted in AD, including the content, sequence and structure of sessions. The existing *Cogtrain* app^20^ will be upgraded to include 4-6 games, with graded difficulty levels, reminders and rewards, and back-end data recording.

#### Treatment As Usual Group (TAU)

Treatment As Usual Group (TAU) will include regular follow-ups with the multidisciplinary team, and medications as prescribed (no changes in dose or duration during the course of the trial), psychoeducation (regarding anxiety, lifestyle modifications, arousal reduction) and rescue protocols in case of crisis.

#### Sample Size

Sample size was calculated based on the previous study.^28^ A total of 100 participants will be recruited across two sites (repeated measures between-group design with two time points, pre- and post-intervention, for primary outcome measure of Hamilton Anxiety Rating Scale (HAM-A), with a correlation between time r = 0.4, with 5% Type I error and power of 90%; n=38 per group + anticipating 30% dropout, rounded off to 50 in each group).

#### Operational Definition

- Cognitive Control: Cognitive control, also called executive function, comprise the following: inhibition - ability to ignore distraction; working memory -ability to hold information in mind and manipulate it; cognitive flexibility - ability to flexibly switch perspectives, focus of attention, or response mapping.^29^
- Completers: The individuals who will attend 80% of the training sessions and/or homework tasks will be qualified as completers.
- Dropouts: Patients who will complete baseline assessments and initiate therapy but later stop reporting for sessions or communicate their unwillingness to attend sessions or miss three consecutive sessions will be considered as dropouts.

#### Implementation Plan

Random numbers will be generated with a 1:1 ratio using permuted block randomization with varying block sizes, stratified by treating centers and concealed in a sealedenvelope, till assignment. The randomization and allocation of participants to two treatment arms shall be done by team member not involved in delivering the intervention. Participants, recruited from the outpatient services of site A and site B (masked for review), who meet criteria and provide consent will undergo baseline assessments and then be randomized to either arm, as indicated in Fig 2. Participants and therapists cannot be blinded to treatment due to the nature of interventions; however, post-intervention clinician ratings will be conducted by a team member blind to pre-intervention scores. Parallel forms will be used for neuropsychological measures where feasible, to minimize practice effects. The intervention will be delivered by qualified clinical psychologists at both sites, under the supervision of consultant clinical psychologists with 15-30 years of experience.

#### Ethics Review

The study proposal has been approved by the Institute Ethics Committee (IEC) at both sites. The trial has been registered with the Clinical Trial Registry of India (masked for review). Participants will be recruited after obtaining written informed consent. Voluntary participation, confidentiality and privacy during data collection and storage will be ensured. Project staff delivering the intervention will monitor clinical risk, supervised by clinical psychologists/psychiatrists with 10-30 years of experience. In case of lack of improvement, or worsening, appropriate management and referral to the treating team/emergency services will be initiated at the respective study sites. In case changes in treatment are indicated, the participant will be considered a drop-out to maintain the integrity of the trial. Data of dropouts will be used till the time of dropout.

Session management will be enabled on the app to minimize the possibility of overuse, and session data will be carefully monitored to identify and appropriately treat participants at risk of technology addiction. A Data and Safety Monitoring Board (DSMB), independent of the funding agency and with no financial, scientific or other conflict of interest with the study, will be constituted to monitor the trial, comprising statisticians and mental health professionals with relevant expertise. The DSMB shall meet annually to ensure smooth and timely recruitment and to assess participant benefit versus risk, reporting of serious adverse events if any, and to advise on interim analyses if needed.

Data will be stored de-identified in password protected systems and data repositories as per ICMR guidelines. As per IT Act (2000) and The Digital Personal Data Protection Act (2023), Sensitive Personal Data or Information (2011), anonymization and encryption, role-based access, secure cloud infrastructure, identity verification and authentication will be ensured.^30–32^ A secure, researcher-managed registration platform and cloud-based backend infrastructure have been established to control user authentication, protect data privacy, and enable logging, and automated backups.

#### Quality Assurance

Assessments and interventions will be delivered by trained clinical psychologists under supervision. Harmonization exercises will be undertaken under the supervision of a clinical neuropsychologist with over 15 years of experience, for administration and scoring of the neuropsychological measures and ICCT techniques to ensure uniformity across sites. Inter-rater reliability exercises will be conducted for clinician ratings, under the supervision of consultant psychiatrists with 10-20 years of experience. In order to minimize practice effects for cognitive measures on pre-post assessments, parallel forms / computerized dynamic task adjustment will be used. Post-assessment clinical ratings will be performed by a rater blind to pre-assessment scores. Further, to ensure fidelity for ICCT, the therapists will complete a brief checklist after each session, to record treatment stage, mode of the session, punctuality, homework, missed sessions, and any deviation from ICCT techniques.

Therapists will undergo training on the app related to participant onboarding, dashboard use, data export, and registration issues. Tutorials and FAQs will be prepared for participants’ app use and troubleshooting. User authentication and registration to ensure that the app is not used on devices other than by study participants. Standard text reminders for appointments will be sent to participants of both groups. For the ICCT group, participants will receive app notifications to complete homework. In addition, participants will be provided with handouts, information and homework sheets and daily log to monitor their progress between sessions. During the follow-up period, participants of both groups will receive once-monthly phone calls to promote retention.

### Data collection and statistical analysis plan

#### Measures

Table 1 provides a list of screening, clinical measures, cognitive measures, socio-occupational measures, and neuroimaging measures. All the measures will be performed at baseline and post-intervention. At midline and follow-up, only clinical, self-report cognitive, and socio-occupational measures will be performed.

#### Screening measures

Besides socio-demographic, clinical and treatment information, diagnosis will be established using the DIAMOND;^27^ and right-handedness using the Edinburgh Handedness Inventory – Short Form.^33^

#### Clinical measures

The primary outcome measure will be the Hamilton Anxiety Rating Scale (HAM-A), a 14-item, clinician-rated scale of anxiety severity.^34^ Secondary outcomes include Montgomery-Asberg Depression Rating Scale (MADRS),^35^ 10-item clinician-rated scale of depression severity and Clinical Global Impression-Severity (CGI-S), a clinician-rating of overall severity of illness on a 7-point scale.^36^

#### Socio-occupational Measure

Work and Social Adjustment Scale (WSAS) is a 5-item self-report global measure to assess functional impairment caused by a psychiatric disorder.^37^

#### Cognitive Measures

Cognitive measures will include standardized neuropsychological measures previously used in Indian samples - Digit Span for verbal working memory and Spatial Span for visuo-spatial working memory;^38^ Colour Trails Test for focused attention and flexibility;^39^ Object Alternation-Probabilistic Reversal Learning (OAT-PRL) for flexibility;^40,41^ Stroop Test for inhibitory control;^42^ Zoo Map Test ^43^ for planning; Logical Memory for verbal learning and memory;^38^ Block Design Test as an estimate of intelligence;^44^ Cognitive Assessment Instrument for Obsessions and Compulsions (CAIOC-13)^45,46^ for self-reported cognitive difficulties; Metacognitive Awareness and Regulation Scale (MARS)^5^ for metacognitive awareness of cognitive abilities in adults with psychiatric disorders. Due to non-availability of standardized Stroop stimuli in Indian languages, a computerized Stroop test in five Indian languages (Hindi, Kannada, Tamil, Telugu, Malayalam) will be newly developed for the study, based on existing original^42^ and computerized versions.^47^ In addition, other self-report measures (WSAS, MARS and CAOIC - 13) will be translated to Indian languages.

Additionally, functional Magnetic Resonance Imaging (fMRI) during Stop-Signal Task (SST)^41^ will be used to investigate changes in the CC circuit pre- and post-intervention only in site A. For fMRI acquisition, echo planar imaging (EPI) images will be acquired using the following parameters: (TR= 2200ms;TE=28 ms;flip angle =80; Slice thickness=3mm; Slice order: Ascending; Slice number = 44; Gap = 10%; Matrix = 64*64*64mm3, FOV =211*211, voxel=3.3 mm. In the resting state dynamics will be 275 scans with 5 dummies lasting for 10.20 minutes. The structural MRI will include T1 for registration, followed by FLAIR and T2 to check for incidental findings. The task lasts for 17 minutes with extra dynamics to accommodate both slow and fast performers.

### Statistical Analysis

Descriptive statistics will be calculated to examine the demographic details of the participants. Change scores will be calculated to examine pre-and post-intervention differences, and response to treatment on the primary and secondary outcome measures. Normality of the distribution will be assessed, and appropriate parametric/non-parametric statistical tests will be chosen viz., t-test/ chi-square test will be used to identify potential confounders. Between-group comparison of pre-, mid- and post-intervention scores will be done using Repeated Measures ANOVA (RMANOVA) for the outcome measures of HAM-A, CGI, MADRS, WSAS, and neuropsychological measures. Use of mixed effects models will be considered if feasible. Missing observations will be handled using suitable Intent-to-Treat analyses based on missing patterns.

Correction in statistical significance (p value) for multiple comparison and testing would be done based on Bonferroni correction. Sensitivity analyses with clinical and demographic variables, and dropouts will be undertaken.

**Timeline :**Figure 3 depicts the proposed timeline of the study

## Discussion

Despite significant progress in treatment of ADs, there is an ongoing search for effective, economical and scalable interventions for resource-constrained settings. Cognitive dysfunction contributes to significant socio-occupational disability in AD - affecting the individual, family, community, and economy, and is not fully addressed by existing treatments. In this context, novel interventions for the unaddressed cognitive dysfunction are critical. Earlier studies have emphasized attentional strategies and emotional working memory in ADs but are characterized by limited generalization of improvements to untrained domains.^10,12–14^ ICCT attempts to address this gap through process-specific training in multiple domains, with exercises focusing on metacognitive training and generalization, with dose, duration and frequency guided by evidence-based recommendations. ICCT, with preliminary evidence in other disorders,^16,21,22^ is suitable for adaptation to ADs. Hence, a study protocol following SPIRIT guidelines is presented in the current paper to test the effectiveness of ICCT in a multi-center RCT.^48^

### Expected limitations and strengths

#### Strengths

A randomized controlled design across multiple centers allows for greater generalization across varied settings and socio-cultural milieu. The inclusion of follow-up assessments will contribute to an understanding of durability of changes, and continued transfer to broader domains of functioning. To the best of our knowledge, this will be the first study examining neural mechanisms of CT using neuroimaging in ADs. Smartphone apps include several advantages - user-friendliness and flexibility for diverse user demographics, standardized intervention delivery, accurate data recording, adaptive difficulty levels, scalability and access to care. The study has important implications for the National Mental Health Program (NMHP) through potential to enhance full functional recovery in ADs and reduce mental health system burden and treatment gaps in the context of resource-limited settings such as India.

#### Limitations

The intervention is being tested in a randomized controlled trial, nevertheless, the problem of blinding and placebo in psychological intervention research has been repeatedly highlighted.^11^ At present, ICCT is suggested as an adjunct to TAU, and future studies may need to test its efficacy as a stand-alone treatment, applicability to severe anxiety, and non-inferiority to other evidence-based psychological therapies such as cognitive behaviour therapy. Factors relating to motivation, engagement, emotional state, and individual differences in reward sensitivity are known to moderate treatment outcomes in cognitive training,^49^ and are likely to impact the proposed study.

User engagement with the app is difficult to ensure, although the smartphone app is expected to record engagement metrics, potentially available as a covariate in statistical analysis. Finally, the long-term stability of gains is not addressed in this study.

#### Expected Outcomes

The study is expected to bridge the knowledge gap on ADs, an under-researched, yet common disorder. Based on previous trials, ICCT is expected to demonstrate improvements in cognitive functions. Since the trained parameters are core transdiagnostic constructs, generalization is expected across neuropsychological domains, clinical symptoms, socio-occupational functioning, and underlying neural circuits of cognitive control; along with good acceptability and adherence, and maintenance of gains at follow-up. This study may also open up avenues to explore diverse clinical conditions such as substance use disorders, attention deficit hyperactivity disorder, mood disorders; and adolescent and older populations. The multimodal assessments, including neurocognitive, metacognitive, neuroimaging, clinical profile, and socio-occupational functioning measures, may shed light on predictors of response to cognitive training, moderator variables, possible mechanisms of action, and necessary and sufficient components of the intervention.

## Conclusions

This paper presents the study protocol for testing effectiveness of ICCT against TAU. Empirical testing of the intervention is expected to bridge the knowledge gap on treatments available for ADs, a common yet under-researched disorder. Digital delivery through a smartphone app may enhance scalability and access. Further, ICCT may potentially merit extension to other clinical conditions.

## Data Availability

All data produced in the present study are available upon reasonable request to the authors.

## Acknowledgements

The authors thank Dr Ashoo Gover, Dr Neha Dahiya, Dr Ravinder Singh, and Dr Harpreet Singh from the ICMR, Professor V. L. Nimgaonkar, Dr Mary Hawk, and the PRIIIA faculty. The content of this manuscript is solely the responsibility of the authors and does not necessarily represent the official views of NIH or FIC, who had no role in the design and conduct of the study; collection, management, analysis, and interpretation of the data; preparation, review, or approval of the manuscript; and decision to submit the manuscript for publication.

## Statements and Declaration

### Consent to Participate

There are no human participants in this article and informed consent is not required.

### Consent to Publication

Not applicable

### Decalaration of Conflicting Interest

The authors declared no potential conflicts of interest with respect to the research, authorship, and/or publication of this article.

### Data availability

Data will be made available on request

## Reporting guideline (Supplementary online material)

**1.Name:**The research protocol is presented in accordance with the SPIRIT 2013 Statement (Standard Protocol Items: Recommendations for Interventional Trials).

## Declaration Regarding the Use of Generative AI

No part of this article was written or generated by a generative AI tool. The authors take full responsibility for the accuracy, integrity, and originality of the published article.

## Citation Diversity Statement

We are committed to equitable citation practices and have made conscious efforts to include work by authors from diverse genders, geographic regions (including the Global South), career stages, and historically marginalized groups. We aim to support a more inclusive and representative scholarly record.

