## Supplementary material for "Smartphone app-based Integrated Cognitive Control Training for Anxiety Disorders: Study protocol for a Multi-site Randomized Control Trial": De-identified informed consent - ICCT + control.docx

**INFORMED CONSENT FORM FOR PARTICIPANTS**

**Information to Participants:**

*What is this study about?*

Anxiety disorders are a commonly occurring group of psychiatric conditions, including Generalized Anxiety Disorder, Social Anxiety Disorder, Specific Phobia, and Panic Disorder. Anxiety disorders may often affect a person’s cognition, i.e., thinking (concentration, memory, planning etc), resulting in difficulties with studies/work, and everyday functioning. The established treatment methods for anxiety disorders include medications, and cognitive behaviour therapy (CBT). Although both are effective, they do not specifically target difficulties in cognition, which individuals with anxiety disorders may experience. Hence, this study attempts to test a newly developed smartphone app-based intervention, Integrated Cognitive Control Training (ICCT), for improving cognitive functions, along with anxiety symptoms and everyday functioning.

*Why are you being invited to be part of the study?*

You are being invited to participate as one of approximately 100 subjects in a research study which will compare two treatments - ICCT and treatment-as-usual (TAU).

We are carrying out this study to examine the comparative usefulness of ICCT and TAU through assessments before and after the treatments (at 8-12 week interval). Since you have been diagnosed with anxiety disorder and may benefit from these treatments, you are being invited to take part in this study.

*What are we doing in the study?*

Each participant will be allocated to a different treatment (lasting approximately 8 weeks) based on random allocation (that is, you will have an equal chance to be in either of the two groups). Random allocation is a statistical technique that makes the findings more trustworthy.

If you are allocated to ICCT, you will meet a clinical psychologist approximately once a week for 40-60 minutes to discuss and practice methods to improve cognitive control, i.e., the ability to modulate attention, inhibit distractions, and use effective strategies to work towards the goal of a task. ICCT will also involve using a smartphone application about four days per week for approximately 20-30 minutes, to engage in games / mental exercises with specific instructions. ICCT has been tested in other psychiatric conditions and demonstrated improvements in cognition and clinical symptoms in smaller studies.

If you are allocated to TAU, you will receive ‘treatment as usual’ for anxiety disorders, which includes routine follow-ups, psychoeducation about the disorder and/or medications as prescribed by a psychiatrist. Medications for anxiety disorders usually include serotonin reuptake inhibitors (SRIs). Psychoeducation includes education/awareness about the disorder and its symptoms, causes and treatments, and is known to be helpful for recovery. You would have routine follow-ups as per your convenience and need.

Before and after either of the treatments, we will administer some assessments in order to examine the effect of the intervention. The assessments include paper-pencil tests of cognition, as well as clinical symptoms which will be administered individually in a testing room. In addition, your brain activity will be recorded while you lie down in a Magnetic Resonance Imaging (MRI) scanner. This will be done to understand the effects of the treatment on brain areas known to be involved in cognitive control, i.e., the focus of the ICCT treatment. This technique is non-invasive (nothing is inserted into the body) and will not harm you. You will be shown a video of the procedure before, and will also have a trial run before the start, to help prepare you for the procedure and address any concerns you may have.

*What is the duration and frequency of study visits?*

ICCT sessions will be once-weekly, approximately 40-60 minute sessions for 8 weeks, with additional homework practice on the smartphone app (20-30 minutes, four times a week approximately). TAU will involve routine follow-ups as per need, but no more than once weekly. All participants will undergo ‘before-after’ assessments. The assessments, including MRI, may take approximately two-three hours, and will be done twice, at an 8-12 week interval. In addition, you will undergo two brief assessments (approximately 4 weeks after starting, and again 6 months after starting), which may take 5-10 minutes, and can also be done remotely (telephone / online) as you prefer.

The treatment and assessment sessions may also be carried out online if you prefer; but the MRI will require in-person visit to (masked for review). All appointments can be fixed at a time of mutual convenience.

*What happens with the data collected?*

All information obtained from the assessment and treatment sessions will be kept confidential, and will be used only in scientific discussions such as conferences and research publications, without disclosing identity or individual responses. The data will be stored de-identified (assigned a number code, without mentioning name), in password-protected devices/folders for five years, with access only to the investigators. De-identified data may also be shared across the study sites (masked for review) or on data repositories as per funding agency requirements.

*What are the anticipated risks?*

While the assessments, brain imaging, and treatments are unlikely to cause harm to participants, if some specific items on the tests or questionnaires cause any distress, please feel free to let us know, and the distress can be addressed.

For the MRI, safety precautions as per international standards will be adhered to throughout. There may be heightened noise levels; ear muffs will be provided to reduce noise. You will be able to communicate with the researcher if needed during the procedure. For some individuals, anxiety levels may increase slightly while in the scanner, but this is expected to be temporary, and will subside quickly. The researcher is a trained clinical psychologist who can assist in managing anxiety if needed. You can choose to pause or discontinue the scan at any time. All efforts will be made to maximize your comfort during the scan. The information video and trial run can help prepare you for the process, and address your concerns.

In case significant other psychiatric / medical difficulties emerge during assessment or treatment, these will be discussed with you, and you will be referred to the treating team for further intervention if needed.

*What are the anticipated benefits?*

Possible benefits of participating in this study may include improvement of your condition and relief from your symptoms. The study researcher will be monitoring your condition more closely than usual.

While the assessments and neuroimaging are likely to provide useful information, you may or may not benefit personally from this undertaking or the treatments, but knowledge gained from this will be used for the benefit of others. The study findings may help establish newer treatments to improve cognition and clinical symptoms in anxiety disorders.

*Who has reviewed this study?*

The study has been approved and funded by (masked for review), and reviewed and approved by the (masked for review) Ethics Committee.

**Undertaking by the investigator:**

Your participation is voluntary. You may or may not benefit personally from this undertaking, but knowledge gained from this will be used for the benefit of others. You have the right to refuse to take part in the study, without adversely affecting your treatment. You can also withdraw your consent during any part of the study. If you have any doubts or questions during the study, you are free to clarify the same with the investigators (details below).

**Consent:**

“I have been informed about the procedures of the study. I have understood that I have the right to refuse my consent or withdraw it any time during the study without adversely affecting my treatment. I am aware that by subjecting myself to this investigation, I will have to give my time and that these assessments will not interfere with the treatment / services. I have understood that I should not participate in any other study till the completion of this study.

I, …………………………………………., the undersigned, give my consent to be a participant of this study.

Signature of the participant	Signature of the witness

Name and Address	Name and Address

Date:	Date:

Signature of the Investigator

Name and Designation:

Date:	Place:

*Details of investigators*

(masked for review)
