## Supplementary material for "Smartphone app-based Integrated Cognitive Control Training for Anxiety Disorders: Study protocol for a Multi-site Randomized Control Trial": Figures _ 2_IJPM.docx

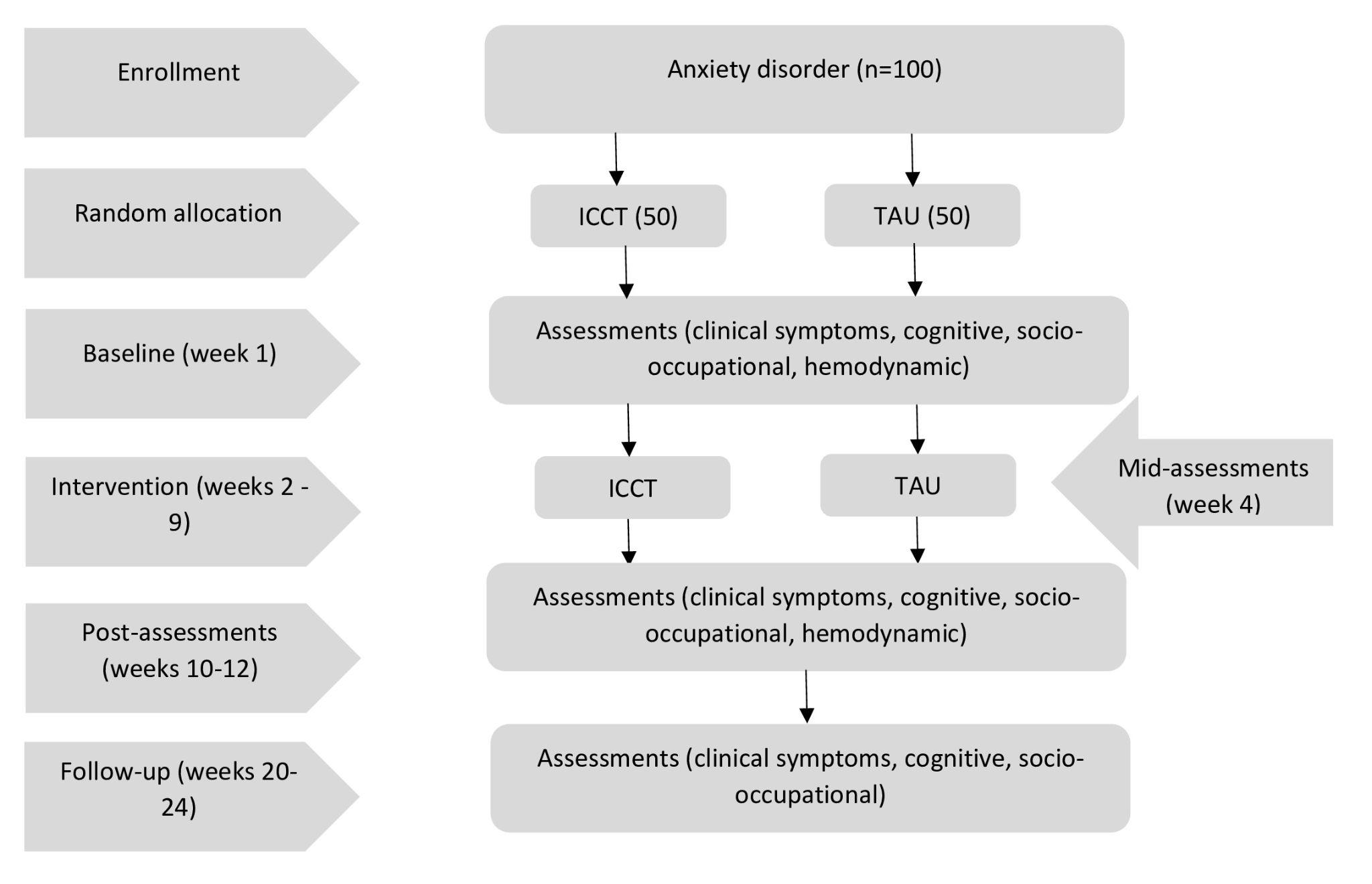

**Figure 2 - CONSORT Flow Diagram depicting participant flow**

*Clinical measures: Hamilton Anxiety Rating Scale (HAM-A), Montgomery-Åsberg Depression Rating Scale (MADRS),Clinical Global Impression (CGI); Cognitive measures: Digit Span, Spatial Span, Colour Trait Test, Stroop Test, Zoo Map Test, Logical Memory, Block Design, Object Alternation Test (OAT), Cognitive Assessment Instrument Obsessions & Compulsions (CAIOC - 13), Metacognitive Awareness and Regulation Scale (MARS); Socio-occupational measure: Work and Social Adjustment Scale (WSAS); Hemodynamic measure: Magnetic Resonance Imaging (MRI), Task based MRI.*
