## Supplementary material for "Smartphone app-based Integrated Cognitive Control Training for Anxiety Disorders: Study protocol for a Multi-site Randomized Control Trial": Table - IJPM.docx

**Table 1- Outcome measures**

| Domain | Measure | Instrument | T0 | T1 | T2 | T3 |
| --- | --- | --- | --- | --- | --- | --- |
| Symptom severity | Anxiety severity | HAM-A | ✔ | ✔ | ✔ | ✔ |
|  | Depression | MADRS | ✔ | ✔ | ✔ | ✔ |
|  | Illness severity | CGI | ✔ | ✔ | ✔ | ✔ |
| Cognitive functions (neuropsychological measures) | Verbal working memory | Digit Span* | ✔ | 🗶 | ✔ | 🗶 |
|  | Visuo-spatial working memory | Spatial Span* | ✔ | 🗶 | ✔ | 🗶 |
|  | Focused attention, flexibility | Colour Trait Test | ✔ | 🗶 | ✔ | 🗶 |
|  | Flexibility | OAT# | ✔ | 🗶 | ✔ | 🗶 |
|  | Response inhibition | Stroop Test# | ✔ | 🗶 | ✔ | 🗶 |
|  | Planning ability | Zoo Map Test | ✔ | 🗶 | ✔ | 🗶 |
|  | Verbal learning & memory | Logical Memory | ✔ | 🗶 | ✔ | 🗶 |
|  | Intelligence | Block Design | ✔ | 🗶 | ✔ | 🗶 |
|  | Perceived Cognitive difficulties | CAIOC-13 | ✔ | ✔ | ✔ | ✔ |
|  | Metacognitive awareness | MARS | ✔ | 🗶 | ✔ | 🗶 |
| Socio-occupational | Functional impairment | WSAS | ✔ | ✔ | ✔ | ✔ |
| Hemodynamic measure |  | Resting - state MRI #  Task Based MRI # | ✔ | 🗶 | ✔ | 🗶 |

*HAM-A - Hamilton Anxiety Rating Scale; MADRS - Montgomery-Åsberg Depression Rating Scale; CGI - Clinical Global Impression (CGI); OAT - Object Alternation Test; CAIOC - 13 - Cognitive Assessment Instrument Obsessions & Compulsions; MARS - Metacognitive Awareness and Regulation Scale; WSAS - Work and Social Adjustment Scale; MRI - Magnetic Resonance Imaging*

*T0 - baseline (Week 1), T1 - Mid-intervention (Week 4), T2 - Post-intervention (Week 10-12), T3 (Week 20-24)
* Parallel forms for different timepoints
\# Computerised tasks with random presentation/adaptive difficulty levels*
