## Supplementary figures and images for "Smartphone app-based Integrated Cognitive Control Training for Anxiety Disorders: Study protocol for a Multi-site Randomized Control Trial"

### Figures _ 1_IJPM.docx

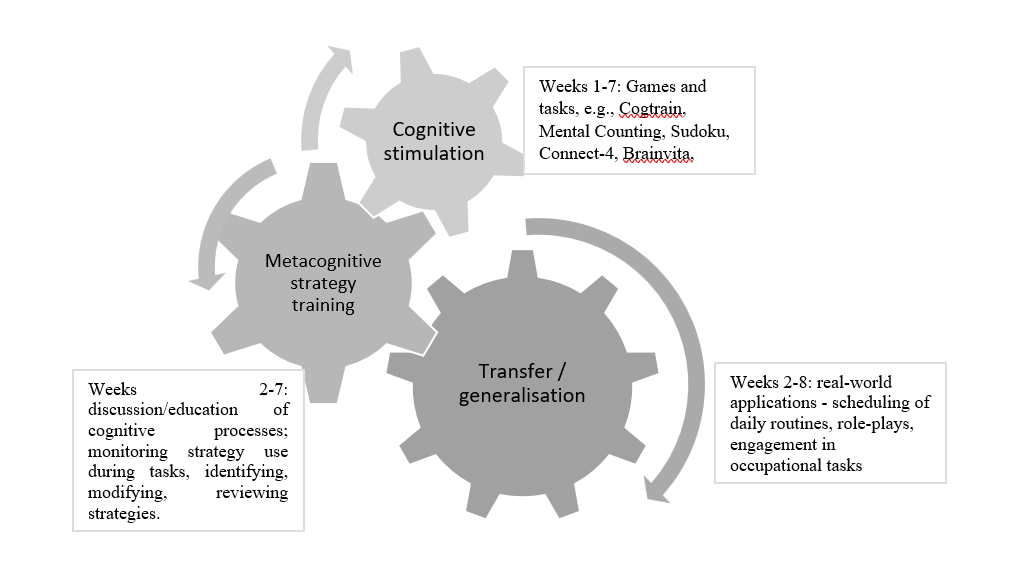

**Figure 1 - Components of Integrated Cognitive Control Training**

### Figures _ 3_IJPM.docx

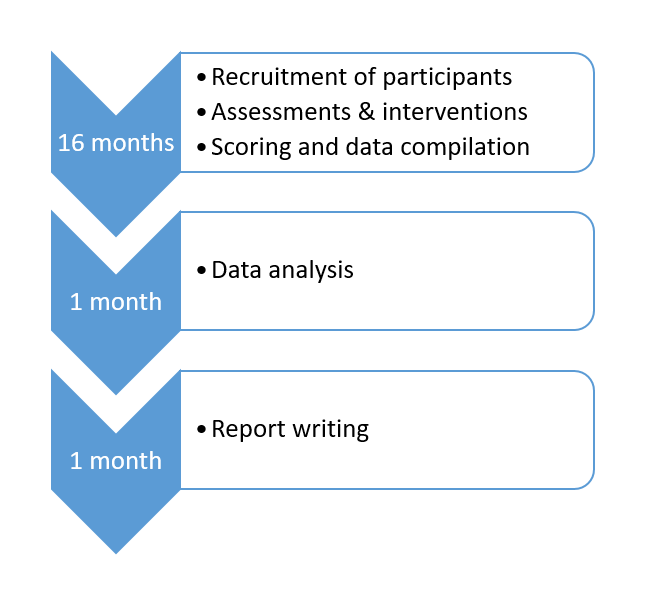

**Figure 3 - Proposed timeline of the protocol**
